# Human genetic evidence supports MAP3K15 inhibition as a therapeutic strategy for diabetes

**DOI:** 10.1101/2021.11.14.21266328

**Authors:** Abhishek Nag, Ryan S. Dhindsa, Andrew R. Harper, Dimitrios Vitsios, Andrea Ahnmark, Bilada Bilican, Katja Madeyski-Bengtson, Bader Zarrouki, Quanli Wang, Katherine Smith, Dave Smith, Benjamin Challis, Dirk S. Paul, Mohammad Bohlooly-Y, Mike Snowden, David Baker, Regina Fritsche-Danielson, Menelas N. Pangalos, Slavé Petrovski

**Affiliations:** Centre for Genomics Research, Discovery Sciences, BioPharmaceuticals R&D, AstraZeneca, Cambridge, UK; Centre for Genomics Research, Discovery Sciences, BioPharmaceuticals R&D, AstraZeneca, Waltham, USA; Bioscience Metabolism, Early CVRM, BioPharmaceuticals R&D, AstraZeneca, Gothenburg, Sweden; Discovery Biology, Discovery Sciences, BioPharmaceuticals R&D, AstraZeneca, Gothenburg, Sweden; Emerging Innovations, Discovery Sciences, BioPharmaceuticals R&D, AstraZeneca, Cambridge, UK; Translational Science and Experimental Medicine, Early CVRM, BioPharmaceuticals R&D, AstraZeneca, Cambridge, UK; Discovery Sciences, BioPharmaceuticals R&D, AstraZeneca, Cambridge, UK; Bioscience Metabolism, Early CVRM, BioPharmaceuticals R&D, AstraZeneca, Cambridge, UK; Early CVRM, BioPharmaceuticals R&D, AstraZeneca, Gothenburg, Sweden; BioPharmaceuticals R&D, AstraZeneca, Cambridge, UK; Departments of Medicine and Neurology, University of Melbourne, Royal Melbourne Hospital, Melbourne, Victoria, Australia

## Abstract

Diabetes mellitus is a chronic health condition that can result in significant end-organ complications and is estimated to impact at least 8.5% of the global adult population. Here, we performed gene-level collapsing analysis on exome sequences from 454,796 multi-ancestry UK Biobank participants to detect genetic associations with diabetes. Rare non-synonymous variants in *GCK, GIGYF1, HNF1A*, and *HNF4A* were significantly associated (*P*<1×10^-8^) with increased risk of diabetes, whereas rare non-synonymous variants in *MAP3K15* were significantly associated with reduced risk of diabetes. Recessive carriers of rare non-synonymous variants in the X chromosome gene *MAP3K15* had a 30% reduced risk of diabetes (OR=0.70, 95% CI: [0.62,0.79], *P*=5.7×10^-10^), along with reduced blood glucose (beta=-0.13, 95% CI: [-0.15,-0.10], *P*=5.5×10^-18^) and reduced glycosylated haemoglobin levels (beta=-0.14, 95% CI: [-0.16,-0.11], *P*=1.1×10^-24^). Hemizygous males carrying protein-truncating variants (PTVs) in *MAP3K15* demonstrated a 40% reduced risk of diabetes (OR=0.60, 95% CI: [0.45,0.81], *P=*0.0007). These findings were independently replicated in FinnGen, with a *MAP3K15* PTV associating with decreased risk of both type 1 diabetes (T1DM) and type 2 diabetes (T2DM) (p<0.05). The effect of *MAP3K15* loss on diabetes was independent of body mass index, suggesting its protective effect is unlikely to be mediated via the insulin resistance pathway. Tissue expression profile of *MAP3K15* indicates a possible involvement of pancreatic islet cell or stress response pathways. No safety concerns were identified among heterozygous or recessive *MAP3K15* PTV carriers across over 15,719 studied endpoints in the UK Biobank. Human population genetic evidence supports *MAP3K15* inhibition as a novel therapeutic target for diabetes.

## Introduction

Diabetes mellitus is a worldwide health concern projected to affect 700 million people by 2045^1^. It is currently the leading cause of micro- and macrovascular disease, including kidney failure, blindness, heart disease, and lower limb amputations^2^. Characterized by elevated levels of blood glucose, diabetes mellitus is generally categorised into type 1 diabetes mellitus (T1DM), type 2 diabetes mellitus (T2DM), and other rarer forms. T1DM is caused by autoimmune destruction of insulin-producing pancreatic β-cells, while T2DM is primarily caused by peripheral insulin resistance. Both types of diabetes eventually lead to progressive loss of pancreatic β-cells and deficient insulin secretion.

Genome wide association studies (GWAS) have implicated over 60 loci in T1DM^3^ and many hundreds of loci in T2DM^4,5^. Except for a few loci that map to protein-coding regions (e.g., *PAM*)^4^, the majority reside in non-coding regions of the genome, making it challenging to map the candidate gene and characterise the underlying causal biology. The growing availability of whole-exome sequences in large population-scale biobanks offers unprecedented opportunities to identify protein-coding variants that have demonstrably large effects on human traits and thus potentially constitute more clinically efficacious target opportunities^6^. Identifying loss-of-function variants that protect against disease is of particular interest since these discoveries can provide direct human-validated therapeutic targets^7–9^.

Here, we report a multi-ancestry exome-sequencing association study for diabetes in 412,394 exomes from the UK Biobank (UKB). Using our previously described gene-level collapsing framework^10^, we identified that recessive loss of the X chromosome gene *MAP3K15* was associated with 40% reduced risk of developing diabetes and decreased circulating glucose and haemoglobin A1c levels. The findings were replicated in the FinnGen study, with a PTV in *MAP3K15* associating with decreased risk of both T1DM and T2DM. Furthermore, the loss of *MAP3K15* was not associated with any apparent on-target adverse phenotypes in a phenome-wide assessment of 15,719 clinical endpoints, supporting *MAP3K15* as a potentially safe target for selective therapeutic inhibition.

## Results

### Cohort characteristics and study design

We processed exome sequences from 454,796 UKB participants through our previously described cloud-based pipeline^10^. Through stringent quality control, we removed samples with low sequencing quality, low depth of coverage, and from closely related individuals (**Methods**). For this study, we focused on 90 binary clinical phenotypes related to T1DM and T2DM available in the UKB (**Supplementary Table 1A**). In total, there were 39,044 cases that mapped to at least one of the diabetes-related clinical phenotypes, including 35,035 of European ancestry, 2,262 of South Asian ancestry, 249 of East Asian ancestry, and 1,498 of African ancestry. Measurements for quantitative traits related to diabetes, including blood glucose, glycosylated haemoglobin (HbA1c), and body mass index (BMI), were also available for participants (**Supplementary Table 1B**).

We employed a gene-level collapsing framework to test the aggregate effect of rare non-synonymous variants in each gene (N=18,762) against each of the diabetes-related clinical phenotypes (**Methods**). Each gene-phenotype combination was tested under 10 non-synonymous collapsing models (including one recessive model) to evaluate a range of genetic architectures, as previously described^10^ (**Supplementary Table 2**). We performed two versions of the collapsing analysis: one restricted to individuals of European ancestry and the other a pan-ancestry analysis, as previously described^10^ (**Methods**). No inflation of test statistics was observed in the gene-level collapsing analysis for the 90 diabetes-related clinical phenotypes that were tested (median genomic inflation lambda across all models = 1.01).

### Rare-variant collapsing analysis

We identified four protein-coding genes significantly associated (*P*<1×10^-8^) with at least one diabetes-related clinical phenotype in the European-only analysis (**Figure 1A**; **Table 1; Supplementary Table 3)**. Rare non-synonymous variants in *GCK, GIGYF1*, and *HNF1A* were associated with increased risk of diabetes, whereas rare non-synonymous variants in *MAP3K15* were associated with reduced risk of diabetes. In the pan-ancestry analysis, all four of these genes maintained statistical significance; one additional gene, *HNF4A*, that was associated with increased risk of diabetes, achieved significance in the pan-ancestry analysis (**Supplementary Table 4**).

**Figure 1.**
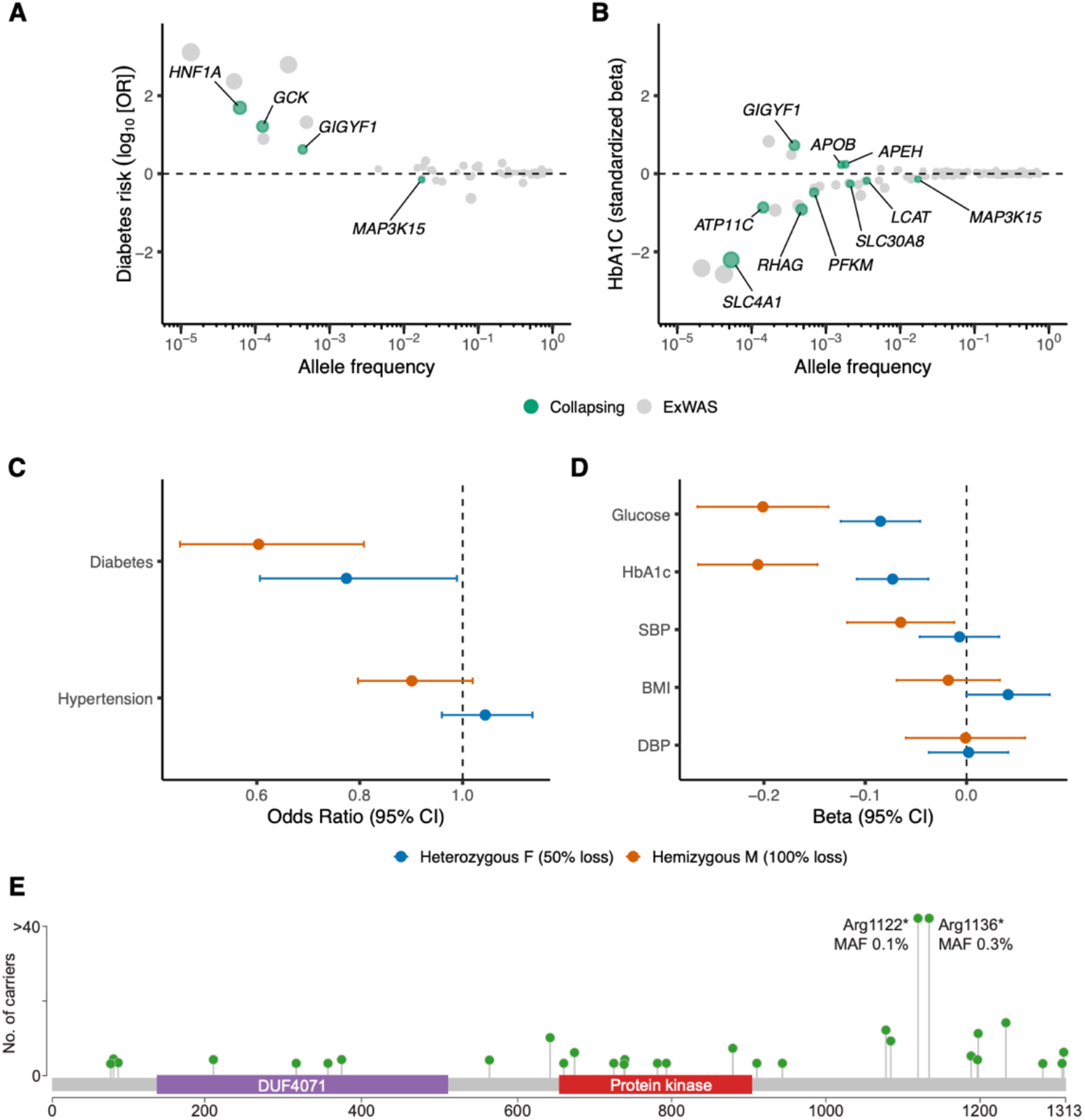
Genetic associations with diabetes and related traits among the European ancestry participants in the UK Biobank. **(A)** Odds ratios and allele frequencies of gene-level (collapsing) and variant-level (ExWAS) associations (p<1×10^-8^) with diabetes diagnoses. **(B)** Effect sizes and allele frequencies of gene-level (collapsing) and variant-level (ExWAS) associations (p<1×10^-8^) with haemoglobin A1c (HbA1c). Allele frequencies on the x-axis are plotted on a log10 scale in both (A) and (B). **(C)** Odds ratios of partial loss (i.e., heterozygous female PTV carriers) and complete loss (i.e., hemizygous male PTV carriers) of *MAP3K15* for diabetes and hypertension diagnoses. **(D)** Effect sizes of partial loss (i.e., heterozygous female PTV carriers) and complete loss (i.e., hemizygous male PTV carriers) of *MAP3K15* for various cardiovascular and metabolic traits related to diabetes. (DBP = Diastolic Blood Pressure, SBP = Systolic Blood Pressure, BMI = Body Mass Index) (**E**) Lollipop plot depicting *MAP3K15* PTVs (stop gain and frameshift variants) observed among hemizygous males of European ancestry. Essential splice variants were not included. The two most frequent PTVs have been annotated with allele frequencies from participants of European ancestry. There were no carriers in common to the two PTVs among the European ancestry males. The y-axis is capped at 40.

**Table 1:** Genes significantly associated with at least one diabetes-related clinical phenotype in the gene-level collapsing analysis among European ancestry participants in the UK Biobank. Genes across the exome were tested under 10 different non-synonymous collapsing models with 90 diabetes-related clinical phenotypes available in the UK Biobank. The most significant diabetes-related clinical phenotype and the corresponding association statistics have been provided for the four genes that were significantly associated (p<1×10^-8^) with at least one diabetes-related clinical phenotype among European ancestry participants. (Chr=Chromosome; QV=Qualifying Variant; OR=Odds Ratio, CI=Confidence Intervals)

| Gene Name | Chr | UKB diabetes-related clinical phenotype | Most significant collapsing model | Frequency of QV carriers |  | OR [95% CI] | P |
| --- | --- | --- | --- | --- | --- | --- | --- |
|  |  |  |  | Cases | Controls |  |  |
| GCK | 7 | 2443#Diabetes diagnosed by doctor | URmtr | 0.25% | 0.04% | 6.75 [4.80,9.50] | 3.14x10<sup>-21</sup> |
| GIGYF1 | 7 | Union#E11#E11 Non-insulin-dependent diabetes mellitus | ptv | 0.13% | 0.03% | 4.00 [2.74,5.84] | 9.98x10<sup>-11</sup> |
| HNF1A | 12 | Source of report of E14 (unspecified diabetes mellitus) | ptv5pcnt | 0.07% | 0.00% | 39.18 [11.06,138.86] | 1.39x10<sup>-10</sup> |
| MAP3K15 | X | Union#E14#E14 Unspecified diabetes mellitus | rec | 1.25% | 1.77% | 0.70 [0.62,0.79] | 5.00x10<sup>-9</sup> |

In our phenome-wide association study of 269,171 European UKB participants^10^, the recessive collapsing model, which includes homozygous, hemizygous, and putative compound heterozygous carriers of rare non-synonymous variants, identified unequivocal associations (*P*<1×10^-8^) between *MAP3K15* and reduced haemoglobin A1C (HbA1c) and glucose levels^10^, accompanied by a suggestive association between *MAP3K15* and decreased risk of diabetes. Consistent with this, a more recent study of 454,787 UKB participants^11^ also found suggestive association between *MAP3K15* and T2DM (OR = 0.85, *P =* 2.8×10^-6^) under an additive genetic model. Among common variant literature, an intronic variant in *MAP3K15* was one of over 300 novel loci reported in a large trans-ethnic GWAS of T2DM^5^ (OR = 1.14, *P =* 1.4×10^-8^). In this current study, with an increased sample size of 394,695 European participants, the association between *MAP3K15* and diabetes reached study-wide significance (p<1×10^-8^) in the recessive model (OR = 0.70; 95% CI: [0.62,0.79], *P* = 5.0×10^-9^). Consistent with our prior findings, the *MAP3K15* qualifying variant (QV) carriers had significantly lower HbA1c levels (beta = −0.14, 95% CI: [-0.16,-0.11], *P* = 3.1×10^-23^) (**Figure 1B**) and blood glucose levels (beta = −0.13, 95% CI: [-0.16,-0.10], *P* = 2.5×10^-17^). In our pan-ancestry analysis, the associations became more significant between *MAP3K15* recessive variants and diabetes (OR = 0.70, 95% CI: [0.62,0.79], *P =* 5.7×10^-10^), HbA1c (beta = −0.14, 95% CI: [-0.16,-0.11], *P* = 1.1×10^-24^), and blood glucose (beta = −0.13, 95% CI: [-0.15,-0.10], *P* = 5.5×10^-18^). Collectively, our results implicate loss of *MAP3K15* as a protective factor for diabetes.

Most of the protective *MAP3K15* signals emerged for T2DM phenotypes (**Supplementary Table 1A**). To determine if *MAP3K15* loss also protects from T1DM, we defined a T1DM-specific phenotype in the UK Biobank (N = 881 cases) using available diagnostic information (**Methods**). The effect of *MAP3K15* recessive variants on T1DM remained in the protective direction, but the association did not achieve study-wide significance at the current T1DM sample size (OR = 0.52, 95% CI: [0.25,1.09], *P* = 0.09).

### Complete versus partial loss of *MAP3K15*

Given that the *MAP3K15* associations emerged strongest among the recessive model, we next tested whether the effect of *MAP3K15* loss on diabetes could be dose dependent. Since *MAP3K15* resides on chromosome X, hemizygous male PTV carriers are expected to have complete loss of the protein, and heterozygous female carriers are expected to have a 50% loss. Consistent with a dose-dependent effect, we found that hemizygous male carriers of European ancestry (N=1,216) demonstrated a 40% decreased risk of developing diabetes compared to male non-carriers (OR = 0.60, 95% CI: [0.45,0.81], *P =* 7.2×10^-4^) (**Supplementary Table 5**). In comparison, heterozygous female carriers (N = 2,604) had a 23% reduced risk of diabetes compared to female non-carriers (OR = 0.77; 95% CI: [0.61, 0.99], *P* = 0.04) (**Figure 1C**). Decrease in HbA1c levels were also three times greater in hemizygous male carriers (beta = −0.21, 95% CI: [-0.26,-0.15], *P =* 1.2×10^-11^) (**Supplementary Table 5**) than in heterozygous female carriers (beta = −0.07, 95% CI: [-0.11,-0.04], *P =* 5.3×10^-5^) (**Figure 1D**). The significantly stronger effects observed with complete loss of *MAP3K15* as compared to partial (50%) loss (**Figures 1C and 1D**) suggests an additive protective effect of *MAP3K15* loss on diabetes-related traits.

We also found that the contributing PTVs occurred throughout the *MAP3K15* gene sequence (**Figure 1E; Supplementary Figure 1**). Two *MAP3K15* PTVs were relatively more frequent and accounted for 74% of the European-ancestry hemizygous male carriers: Arg1122* (MAF = 0.11%) and Arg1136* (MAF = 0.35%) (**Figure 1E; Supplementary Table 6**). Although proximally close, none of the European-ancestry males carried both PTVs. Consistent with this, the two PTVs were found to be independently associated with reduced risk of diabetes (Arg1122*: OR = 0.33, 95% CI: [0.13,0.80], *P* = 0.02; Arg1136*: OR = 0.60, 95% CI: [0.41,0.88], *P* = 0.01) and lower HbA1c levels (Arg1122*: beta = −0.30, 95% CI: [-0.44,-0.15], *P* = 4.3×10^-5^; Arg1136*: beta = −0.20, 95% CI: [-0.27,-0.12], *P* = 1.3×10^-6^) (**Supplementary Table 7**). When excluding these two PTVs from the collapsing test, carriers of the remaining 38 ultra-rare *MAP3K15* PTVs observed among the European-ancestry males also had significantly reduced HbA1c levels (beta = −0.16, 95% CI: [-0.28,-0.05], *P* = 5.2×10^-3^). Due to reduced sample and, thus, statistical power, the association with reduced diabetes risk (OR = 0.84, 95% CI: [0.50,1.39], *P* = 0.49) did not achieve significance among the remaining ultra-rare PTV carriers (**Supplementary Table 7**).

### Replication analysis

Using summary statistics from the FinnGen study release 5 [N=218,792], we next aimed to replicate the *MAP3K15* findings. The two more frequent *MAP3K15* PTVs (Arg1122* and Arg1136*) were both well-imputed (INFO scores: 0.98 and 0.84, respectively) in the FinnGen dataset. The Arg1122* PTV (rs140104197), which is three times more common in individuals of Finnish descent (MAF = 0.33%) than in UKB Europeans (MAF = 0.11%), was significantly associated with protection from both T1DM (OR = 0.58, *P* = 4.9×10^-4^) and T2DM (OR = 0.82, *P* = 0.04) (**Supplementary Table 8**). Arg1136* (rs148312150) was less frequent in individuals of Finnish descent compared to Europeans in UKB (MAF: 0.16% versus 0.35%), and it individually did not reach statistical significance with any diabetes-related phenotype in FinnGen. There were no other *MAP3K15* PTVs detected in FinnGen.

### *MAP3K15* protective PTV signal is not associated with changes in body mass index or metabolic derangements

Obesity, which can lead to increased insulin resistance, is a strong risk factor for T2DM. To investigate whether the effect of *MAP3K15* on diabetes is mediated via adiposity, we further tested the effect of complete loss of *MAP3K15* adjusting for BMI. The associations between hemizygous *MAP3K15* PTV carrier status and both HbA1c (BMI-unadjusted: beta = −0.21, 95% CI: [-0.15,-0.26], *P =* 1.2×10^-11^; BMI-adjusted: beta = −0.20, 95% CI: [-0.14,-0.26], *P =* 1.1×10^-11^) and diabetes (BMI-unadjusted: OR = 0.60, 95% CI: [0.45,0.81], *P =* 7.2×10^-4^; BMI-adjusted: OR = 0.59, 95% CI: [0.43,0.79], *P =* 5.3×10^-4^) remained consistent after adjusting for BMI, suggesting the protective effect of *MAP3K15* loss on diabetes is unlikely to be mediated via insulin resistance and is likely to benefit individuals irrespective of BMI.

Certain genes that influence diabetes risk can also impact other clinically relevant biomarkers. For example, although PTVs in *GIGYF1* are associated with increased risk of diabetes, they are also associated with reduced low-density lipoprotein cholesterol^12^. We thus tested whether collapsing analyses of *MAP3K15* rare non-synonymous variants associated with any of 168 NMR-based blood metabolite measurements available for approximately 120,000 of the UKB participants. Among the studied metabolites, *MAP3K15* was only associated with reduced glucose (“ptv5pcnt” model, beta = −0.16, 95% CI: [-0.23,-0.10], *P* = 4.4×10^-7^).

### Potential *MAP3K15* inhibition safety liabilities

We observed that approximately 1 in every 150 (0.6%) European-ancestry male participants in the UKB has a lifetime systemic absence of functional *MAP3K15*. Given these individuals are participants in a generally healthy cohort such as the UKB provides considerable support to the tolerability of *MAP3K15* inhibition in humans. This is further supported by this gene’s pLI score of 0.0, which is a measure of the tolerance of a given gene to protein-truncating variants^13^.

However, we sought to systematically evaluate whether therapeutically inhibiting *MAP3K15* could associate with on-target adverse phenotypes. To achieve this, we surveyed associations between non-synonymous variants in *MAP3K15* and 15,719 clinical phenotypes in the UKB, as described previously^10^. We did not observe any significant adverse phenotypic associations (*P*<1×10^-8^) in individuals with *MAP3K15* loss (“ptv”, “ptv5pcnt”, and “rec” collapsing models) in either the individual ancestry or the pan-ancestry analysis. We next tested whether there were any other non-diabetes *MAP3K15* associations at a less conservative p-value threshold (*P*<1×10^-4^). In the European ancestry participants, there were no associations in the “ptv” or “ptv5pcnt” model even at this more liberal p-value cut-off. There were two associations observed in models that included missense variants: hepatomegaly with splenomegaly and diseases of the tongue (**Supplementary Table 9A**).

Finally, as previous animal model studies have highlighted that knockout of *Map3k15* in mice models introduces a hypertensive phenotype^14^, we sought to look at this specific phenotype in greater detail. Among the large European sample, there was no evidence of increased risk to hypertension. Instead, the effect of *MAP3K15* PTVs on human blood pressure-related traits seems to be in the protective direction. The hemizygous *MAP3K15* PTV carriers (i.e., complete loss of *MAP3K15*) showed a modest effect in the protective direction for both hypertension *(‘Union#I10#I10 Essential (primary) hypertension*’: OR = 0.90, 95% CI: [0.80,1.02], *P* = 0.10) and systolic blood pressure (beta = −0.07, 95% CI: [-0.12,-0.01], *P* = 0.01) (**Figures 1C and 1D**; **Supplementary Table 10**). Similarly, among the independent FinnGen cohort, the Finnish-enriched *MAP3K15* PTV (Arg1122* (rs140104197)) that associated strongly with T1DM and T2DM, replicated a modest protective effect on hypertension (OR = 0.84, *P* = 8.5×10^-3^). Expanding our assessment to *MAP3K15* missense variants, the strongest signal for hypertension arises among participants of South Asian ancestry (N=8,078), where the recessive collapsing model showed an association between *MAP3K15* and a hypertension phenotype (*‘41202#BlockI10-I15#I10-I15 Hypertensive diseases’*: OR = 6.33, 95% CI: [3.02,13.28], *P =* 4.8×10^-5^) (**Supplementary Table 9A**). Although not study-wide significant (*P*<1×10^-8^), more importantly, this signal was driven by *MAP3K15* missense variants, with seven hemizygous missense carriers and one hemizygous PTV carrier among affected males of South Asian descent (**Supplementary Table 9B**). A more detailed screen of *MAP3K15* missense variants identified rs56381411 associating with an increased risk of hypertension in FinnGen (MAF = 1.5%, OR = 1.17, *P* = 2.4×10^-7^). Evaluation of the effect of *MAP3K15* missense variants in recessive form in European ancestry participants revealed a nominally significant association with a blood pressure phenotype in the UK Biobank too (*‘Union#R030#R03*.*0 Elevated blood-pressure reading*| *without diagnosis of hypertension*’: OR = 1.44, 95% CI: [1.02,2.03], *P* = 0.05).

Whereas certain missense variants in *MAP3K15* may increase the risk of hypertension, there is no evidence for PTVs in this gene conferring risk. Collectively, these findings suggest a potential *MAP3K15* allelic series, whereby putative gain-of-function missense variants in might increase the risk of hypertension while PTV (putative loss-of-function) alleles protect against hypertension. Further functional characterisation of variation in *MAP3K15* is required to better understand their variable effects on blood pressure- and diabetes-related traits. Altogether, the lack of association between PTVs in *MAP3K15* and any adverse phenotypes suggest that there is a low human safety risk for selective therapeutic inhibition of *MAP3K15*.

### Orthogonal evidence

Because *MAP3K15* appears to be associated with reduced risk of T1DM and T2DM and is not associated with BMI, the data suggests that the protective effect is unlikely to be operating through insulin sensitisation. Physiologically, *MAP3K15* encodes a mitogen-activated protein kinase that is known to play a role in regulating cell stress and apoptotic cell-death^15^. To gain more insight into potential protective mechanisms, we examined the tissue expression profiles of *MAP3K15* in GTEx^16^. *MAP3K15* is most strongly expressed in the adrenal glands and is also expressed at relatively lower levels in the spleen, kidney, pancreas, and pituitary glands (**Figure 2A**). Single-cell expression data from human pancreatic endocrine cells indicate that *MAP3K15* is most strongly expressed in islet cell subpopulations, including α–, β– and δ– cells^17–21^ (**Figure 2B**). Its ubiquitous expression profile in the adrenals could suggest a role in mediating catecholamine biosynthesis or glucocorticoid response^22^. Alternatively, its effect could be mediated through maintenance of endogenous pancreatic islet cells.

**Figure 2.**
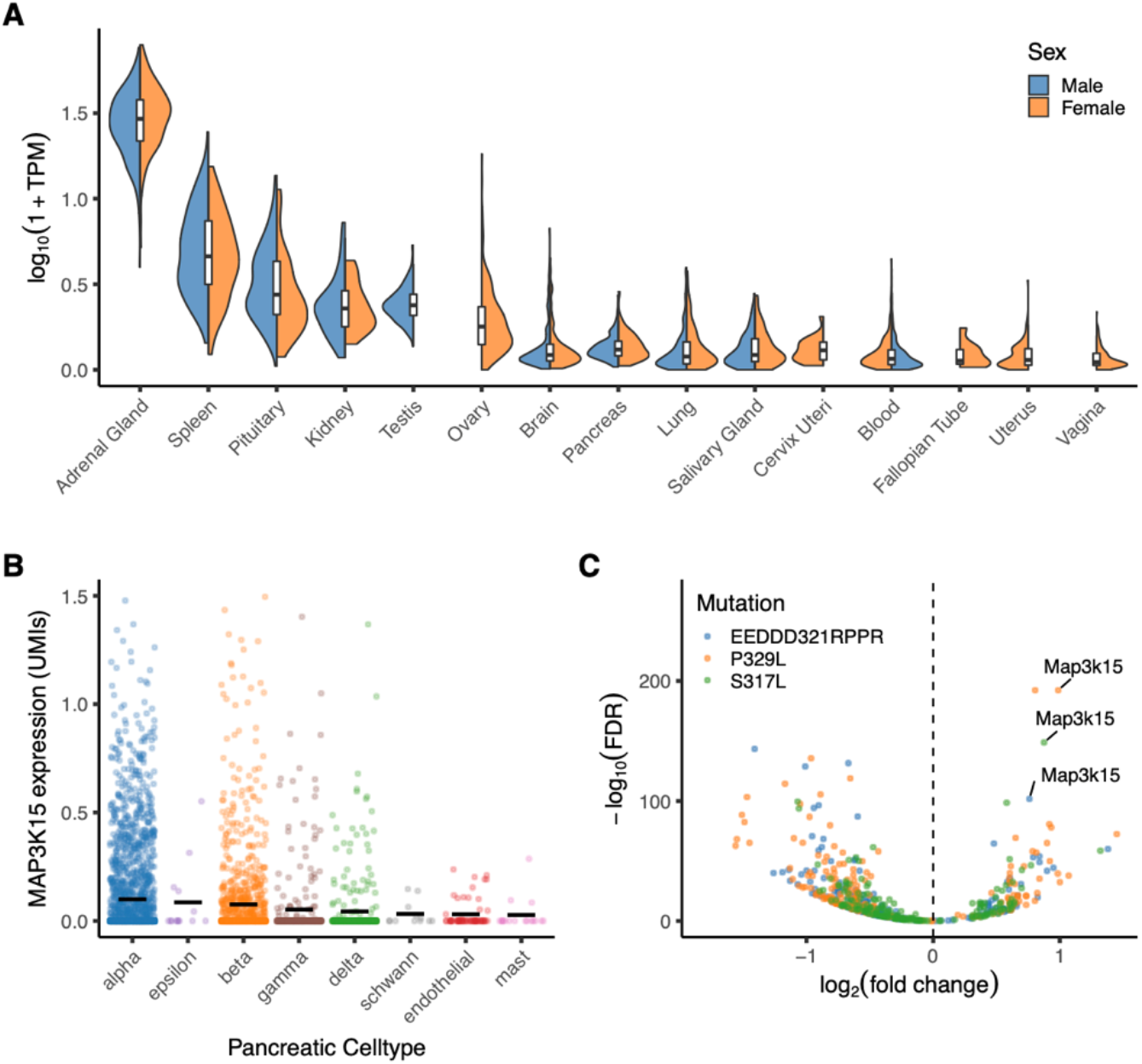
Tissue expression profile of *MAP3K15*. **(A)** Expression of *MAP3K15* in human tissues contained in the GTEx database. TPM = transcripts per million. We only included tissues with a median TPM > 0.1. **(B)** *MAP3K15* expression in major subpopulations of human pancreatic cells derived from a previously published single-cell RNA-sequencing dataset^17–21^. **(C)** Volcano plot depicting differential gene expression in mouse insulinoma cell lines stably expressing three variants in *Nkx6-1:* two MODY-associated variants (P329L and S317L) and a control mutation known to functionally impair Nkx6-1 (EEDD321RPPR)^23^.

To further explore whether *MAP3K15* contributes to the pathophysiology of diabetes in pancreatic cells, we assessed differential gene expression data from a prior study focused on Maturity Onset Diabetes of the Young (MODY)^23^, an early-onset, autosomal dominant form of noninsulin-dependent diabetes. In this study, expression profiling was performed on a mouse insulinoma cell line carrying mutations in the MODY-associated gene *Nkx6-1*. In all three mutations tested, including a positive control mutant known to impair Nkx6-1 as well as two MODY-associated genetic variants, *MAP3K15* was found to be the most significantly upregulated gene (**Figure 2C**). This finding suggests that increased *MAP3K15* activity may mediate the pathophysiology of diabetes, potentially through an increased rate of beta cell loss.

Orthogonal evidence derived from two *in silico* tools, Gene-SCOUT^24^ and phenome-wide Mantis-ML^25^ provide further support for a role of *MAP3K15* in diabetes. Gene-SCOUT provides biomarker fingerprint similarity between any pair of human genes based on UKB exome sequencing cohort statistics using 1,419 quantitative traits^24^. Entering *MAP3K15* as the seed gene in this tool highlights *SLC30A8* as having the most similar human biomarker profile to what is observed for *MAP3K15* (**Figures 3A and 3B; Supplementary Figure 2**). *SLC30A8*, a zinc transporter gene (ZnT8) expressed in pancreatic islet α- and β-cells, is reported to have a protective effect against T2DM potentially via increased glucose responsiveness^26,27^. Mantis-ML, an automated machine-learning framework designed to identify gene-phenotype relationships based on compendium of publicly available disease-specific features (such as tissue expression, preclinical models, genic intolerance, among others), suggests disorders related to impaired glucose homeostasis, including “diazoxide-resistant diffuse hyperinsulinism” and “hyperinsulinemic hypoglycaemia” (**Figure 3C**), among the top 1% of human phenotypes that *MAP3K15* may have a role in **(Supplementary Table 11)**. While mantis-ml does not indicate whether a gene may have a causal versus protective role for a given phenotype, these results converge on *MAP3K15’*s involvement in diabetes-related biology. Both these tools provide diverse and independent support of a biological role for *MAP3K15* in human diabetes.

**Figure 3.**
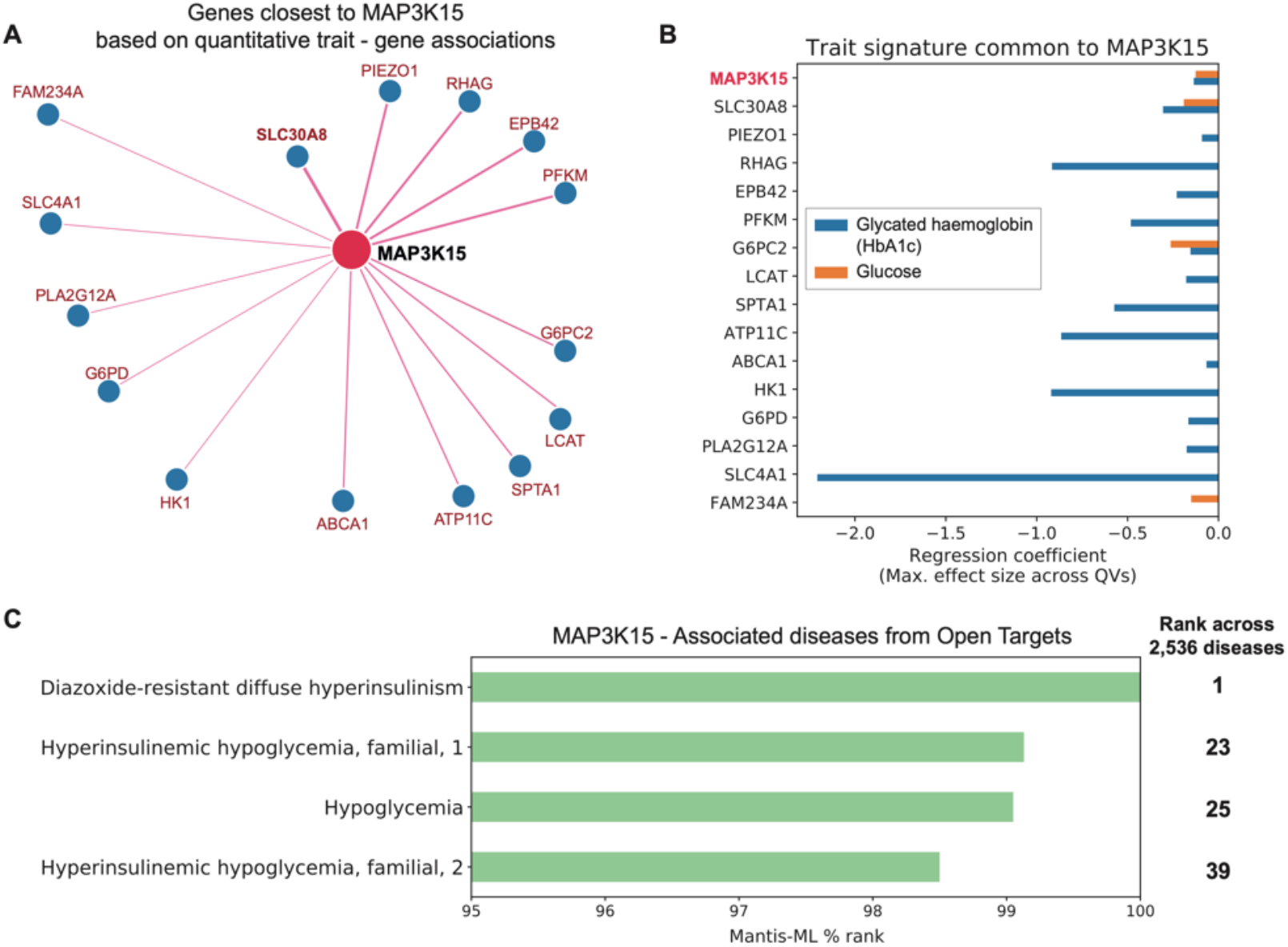
*MAP3K15* quantitative trait and disease signatures. **(A)** Genes with most similar quantitative trait profiles as *MAP3K15* in the UK Biobank, derived from Gene-SCOUT^24^. **(B)** Linear regression coefficients for HbA1c and glucose from collapsing analysis models for genes in panel A (genes are sorted from top to bottom in decreasing order of similarity to *MAP3K15*). QVs = qualifying variants. **(C)** Mantis ML^25^ predictions of *MAP3K15* disease associations.

## Discussion

This exome-sequencing study of 456,796 UKB participants increases our understanding of high-effect size genetic factors involved in both propensity for and protection from diabetes in humans. We found that recessive loss of *MAP3K15* reduces the risk of developing diabetes by approximately 40%. This was supported by the association between recessive loss of *MAP3K15* and decreased HbA1c and blood glucose. Although the protective signal was strongest for T2DM, the effect of *MAP3K15* PTVs was also in the protective direction for T1DM risk in both the UKB and FinnGen. This supports the notion that despite being defined as distinct clinical entities, T1DM and T2DM share some common pathophysiological pathways such as β-cell dysfunction^28,29^.

Crucially, loss-of-function mutations that protect against human disease can act as direct *in vivo* validation of therapeutic targets; thus, *MAP3K15* inhibition could have therapeutic value in both T1DM and T2DM. Given the relatively modest decrease in glucose (0.25 mmol/L) and HbA1c levels (1.36 mmol/mol) that associate with *MAP3K15* loss, one possibility is that the observed protective effect on clinical diabetes is not primarily mediated through glucose / HbA1c reduction. We also found that the protective effect of *MAP3K15* loss is independent of BMI, which might suggest that its effect on diabetes is unrelated to insulin sensitivity, though we note that the relationship between BMI and insulin resistance is correlational. While not currently available for UK Biobank participants, quantitative measures of insulin resistance in *MAP3K15* PTV carriers in future studies could illuminate whether the protective mechanism is indeed unrelated to the insulin resistance pathway. Nonetheless, our results suggest that therapeutic selective inhibition of *MAP3K15* could also benefit patients living with diabetes who are in the low-to-normal BMI range.

Through a phenome-wide association study in the 454,796 human participants, we showed that *MAP3K15* loss is not significantly associated with any phenotypes that would suggest safety concerns due to therapeutic inhibition of this target. Prior work observed that knocking out *Map3k15* in mice led to hypertension^14^. Interestingly, we found that in humans, PTVs appeared to provide a protective effect on hypertension, whereas certain missense variants in *MAP3K15* appeared to increase the risk of hypertension. Collectively, these results not only highlight possible species-specific differences upon loss of *MAP3K15*, but also suggest an allelic series in humans, in which missense variants exert a spectrum of loss- to gain-of-function effects. Future functional characterisation of clinically associated missense variants and PTVs could provide further insights into this potential allelic series.

In our previously published work, *MAP3K15* was one of 15 genes that had unequivocal associations with glucose and/or HbA1c^10^. What sets *MAP3K15* apart from these other genes is that, with the addition of 150,000 more exomes, we also observe a statistically significant reduced risk for diabetes diagnosis, in addition to the biomarker associations. This finding has important implications for the interpretation of genetic biomarker associations. Crucially, not all genetic associations with clinically relevant biomarkers will be related to the pathophysiology of the underlying disease. Here, anchoring biomarker genetic signals with relevant clinical endpoints can help identify those that are more likely to modify the underlying disease. Therapeutically, this suggests that inhibiting *MAP3K15* may target the core pathophysiology of the disease process rather than targeting reduced blood glucose.

The tissue expression profile of *MAP3K15* demonstrates predominant expression in adrenal glands and several islet cell subpopulations, suggesting that *MAP3K15* might be involved in pancreatic islet cell functional maintenance and / or stress response pathways. Dysregulation of stress response in diabetes^30,31^ and the role of *ASK* (*MAP* kinase) family of genes in regulating stress response (e.g., apoptosis, inflammation) to external stimuli^14,15^ offer further support to these mechanisms. These provide important clues regarding the otherwise unknown pathways that mediate the protective effect of *MAP3K15* natural inhibition on diabetes.

## Methods

### Cohorts

Discovery genetic association studies were performed using the 454,796 exomes available in the UK Biobank (UKB) cohort^32^. The UKB is a prospective study of approximately 500,000 participants aged 40–69 years at time of recruitment. Participants were recruited in the UK between 2006 and 2010 and are continuously followed. The average age at recruitment for sequenced individuals was 56.5 years and 54% of the sequenced cohort is of female genetic sex. Participant data include health records that are periodically updated by the UKB, self-reported survey information, linkage to death and cancer registries, collection of urine and blood biomarkers, imaging data, accelerometer data and various other phenotypic end points. All study participants provided informed consent and the UK Biobank has approval from the North-West Multi-centre Research Ethics Committee (MREC; 11/NW/0382).

Replication of the findings in the UKB was performed using the summary statistics from the FinnGen study. The FinnGen cohort (release 5) includes 218,792 individuals from Finland with genotype and health registry data. Phenotypes have been derived from nationwide health registries. Patients and control subjects in FinnGen provided informed consent for biobank research, based on the Finnish Biobank Act. Alternatively, older research cohorts, collected prior the start of FinnGen (in August 2017), were collected based on study-specific consents and later transferred to the Finnish biobanks after approval by Fimea, the National Supervisory Authority for Welfare and Health. Recruitment protocols followed the biobank protocols approved by Fimea. The Coordinating Ethics Committee of the Hospital District of Helsinki and Uusimaa (HUS) approved the FinnGen study protocol Nr HUS/990/2017. The FinnGen study is approved by Finnish Institute for Health and Welfare.

### Phenotypes

We harmonized the UKB phenotype data as previously described^10^. Briefly, we used PEACOK and union mapping to parse binary and quantitative traits included in the February 2020 UKB release (accessed March 27, 2020; UKB application 26041). Here, we considered 90 binary (clinical) phenotypes related to diabetes available in the UKB (**Supplementary Tables 1A**), three quantitative traits related to diabetes (blood glucose, glycosylated haemoglobin and body mass index), and two quantitative traits related to hypertension (systolic blood pressure (SBP) and diastolic blood pressure (DBP)) (**Supplementary Tables 1B**).

Additionally, for a type 1 diabetes (T1DM)-specific analysis, we defined the case population using ICD-9 and ICD-10 codes that captured T1DM diagnoses and the control population by excluding participants with any diabetes diagnoses.

For analyses involving SBP and DBP, we adjusted for commonly prescribed blood pressure medications (**Supplementary Table 12**).

### Genetic data

Exome sequencing data for 454,988 UKB participants were generated at the Regeneron Genetics Center (RGC) as part of a pre-competitive data generation collaboration between AbbVie, Alnylam Pharmaceuticals, AstraZeneca, Biogen, Bristol-Myers Squibb, Pfizer, Regeneron and Takeda with the UKB. Genomic DNA underwent paired-end 75-bp whole-exome sequencing at Regeneron Pharmaceuticals using the IDT xGen v1 capture kit on the NovaSeq6000 platform. Conversion of sequencing data in BCL format to FASTQ format and the assignments of paired-end sequence reads to samples were based on 10-base barcodes, using bcl2fastq v2.19.0. Initial quality control was performed by Regeneron and included sex discordance, contamination, unresolved duplicate sequences and discordance with microarray genotyping data checks.

In FinnGen, genotyping of the samples was done using a ThermoFisher Axiom custom array. In addition to the core GWAS markers (about 500,000), it contains 116,402 coding variants enriched in Finland, 10,800 specific markers for the HLA/KIR region, 14,900 ClinVar variants, 4,600 pharmacogenomic variants and 57,000 selected markers.

### AstraZeneca Centre for Genomics Research (CGR) bioinformatics pipeline

The 454,796 UKB exome sequences were re-processed at AstraZeneca from their unaligned FASTQ state. A custom-built Amazon Web Services (AWS) cloud compute platform running Illumina DRAGEN Bio-IT Platform Germline Pipeline v3.0.7 was used to align the reads to the GRCh38 genome reference and perform single-nucleotide variant (SNV) and insertion and deletion (indel) calling. SNVs and indels were annotated using SnpEFF v4.3^33^ against Ensembl Build 38.92. We further annotated all variants with their genome Aggregation Database (gnomAD) MAFs (gnomAD v2.1.1 mapped to GRCh38)^13^. We also annotated variants using MTR score^34^ to identify if they mapped to genic regions under constraint for missense variants and REVEL scores^35^ for their predicted deleteriousness.

### Additional quality control

To complement the quality control performed by Regeneron Genomics Centre, we passed the UKB exome sequences through our internal bioinformatics pipeline as previously described^10^. Briefly, for UKB, we excluded from our analyses an additional 122 sequences that achieved a VerifyBAMID freemix (measure of DNA contamination) of more than 4%, and an additional 5 sequences where less than 94.5% of the consensus coding sequence (CCDS release 22) achieved a minimum of ten-fold read depth. The cohort was also screened to remove participants that were second-degree relatives or closer (equivalent to kinship coefficient > 0.0884), as determined using the --kinship function in KING v2.2.3^36^. After the above quality control steps, there remained 412,394 unrelated UKB sequences of any genetic ancestry that were available for analyses presented in this study.

### Genetic ancestry

The primary discovery analysis was performed in UKB participants of European ancestry. We used the available exome sequencing data to perform genetic ancestry prediction in PEDDY v0.4.2. We leveraged sequences from the 1,000 Genomes Project as population references^37^ for ancestry estimation. 394,695 (93%) of the 422,488 unrelated UKB participants – that had European ancestry prediction >0.99 and were within 4 SD of the means for the top four principal components – were selected for the European-ancestry case-control analyses. We also used the PEDDY-derived ancestry predictions to identify non-European ancestry populations that had at least 1,000 individuals with exome sequences to perform pan-ancestry collapsing analyses (see the section ‘Collapsing analyses’). This identified 7,412 African, 2,209 East Asian and 8,078 South Asian UKB participants based on predicted ancestry >0.95 for the respective ancestries.

### Discovery analyses

#### Collapsing analyses

We performed our previously described gene-level collapsing analysis framework^10^ for 90 binary and 5 quantitative traits related to diabetes. We included 10 non-synonymous collapsing models, including 9 dominant and one recessive model, plus an additional synonymous variant model as an empirical negative control (**Supplementary Table 2**). For the dominant collapsing models, the carriers of at least one qualifying variant (QV) in a gene were compared to the non-carriers. In the recessive model, individuals with two copies of QVs either in homozygous or putatively compound heterozygous form were compared to the non-carriers. Hemizygous genotypes for X chromosome genes also qualified for the recessive model.

Using SnpEff annotations, we defined synonymous variants as those annotated as ‘synonymous_variant’. We defined PTVs as variants annotated as exon_loss_variant, frameshift_variant, start_lost, stop_gained, stop_lost, splice_acceptor_variant, splice_donor_variant, gene_fusion, bidirectional_gene_fusion, rare_amino_acid_variant, and transcript_ablation. We defined missense as: missense_variant_splice_region_variant, and missense_variant. Non-Synonymous variants included: exon_loss_variant, frameshift_variant, start_lost, stop_gained, stop_lost, splice_acceptor_variant, splice_donor_variant, gene_fusion, bidirectional_gene_fusion, rare_amino_acid_variant, transcript_ablation, conservative_inframe_deletion, conservative_inframe_insertion, disruptive_inframe_insertion, disruptive_inframe_deletion, missense_variant_splice_region_variant, missense_variant, and protein_altering_variant.

For binary traits, the difference in the proportion of cases and controls carrying QVs in a gene was tested using a Fisher’s exact two-sided test. For quantitative traits, the difference in mean between the carriers and non-carriers of QVs was determined by fitting a linear regression model, correcting for age, sex and medication intake (for SBP and DBP).

For all models, we applied the following quality control filters: minimum coverage 10X; annotation in CCDS transcripts (release 22; approximately 34 Mb); at most 80% alternate reads in homozygous genotypes; percent of alternate reads in heterozygous variants ≤ 0.25 and ≥ 0.8; binomial test of alternate allele proportion departure from 50% in heterozygous state P > 1 × 10^-6^; GQ ≤ 20; FS ≥ 200 (indels) ≤ 60 (SNVs); MQ ≤ 40; QUAL ≤ 30; read position rank sum score ≤ −2; MQRS ≤ −8; DRAGEN variant status = PASS; the variant site achieved ten-fold coverage in ≥ 25% of gnomAD exomes, and if the variant was observed in gnomAD exomes, the variant achieved exome z-score ≤ −2.0 and exome MQ ≥ 30. We excluded 46 genes that we previously found associated with batch effects^10^.

#### Pan-ancestry collapsing analyses

We performed additional collapsing analysis in each individual non-European ancestral population as described above. For binary traits, we then performed a pan-ancestry analysis using our previously introduced approach^10^ of applying a Cochran-Mantel-Haenszel test to generate combined 2×2×N stratified P-values, with N representing up to all four genetic ancestry groups. For quantitative traits, the pan-ancestry analysis was performed using a linear regression model that included the following covariates: age, sex, categorical ancestry (European, African, East Asian or South Asian), and top five ancestry principal components.

#### Variant-level (ExWAS) analyses

We performed variant-level association tests in addition to the gene-level collapsing analyses for the 90 binary and 5 quantitative traits related to diabetes. We tested 3.3 million variants identified in at least six individuals from the 394,695 predominantly unrelated European ancestry UKB exomes as previously described^10^. In summary, variants were required to pass the following quality control criteria: minimum coverage 10X; percent of alternate reads in heterozygous variants ≥ 0.2; binomial test of alternate allele proportion departure from 50% in heterozygous state P > 1 × 10^-6^; genotype quality score (GQ) ≥ 20; Fisher’s strand bias score (FS) ≤ 200 (indels) ≤ 60 (SNVs); mapping quality score (MQ) ≥ 40; quality score (QUAL) ≥ 30; read position rank sum score (RPRS) ≤ −2; mapping quality rank sum score (MQRS) ≤ −8; DRAGEN variant status = PASS; variant site is not missing (that is, less than 10X coverage) in 10% or more of sequences; the variant did not fail any of the aforementioned quality control in 5% or more of sequences; the variant site achieved tenfold coverage in 30% or more of gnomAD exomes, and if the variant was observed in gnomAD exomes, 50% or more of the time those variant calls passed the gnomAD quality control filters (gnomAD exome AC/AC_raw ≤ 50%). P values were generated adopting a Fisher’s exact two-sided test. Three distinct genetic models were studied for binary traits: allelic (A versus B allele), dominant (AA + AB versus BB) and recessive (AA versus AB + BB), where A denotes the alternative allele and B denotes the reference allele. For quantitative traits, we adopted a linear regression (correcting for age and sex) and replaced the allelic model with a genotypic (AA versus AB versus BB) test.

#### Phenome-wide analysis for *MAP3K15*

We performed a phenome-wide collapsing analysis for *MAP3K15* with 15,719 binary phenotypes for each individual ancestry in the UKB. We harmonized and union mapped these phenotype data as previously described^10^. We included all 11 collapsing models in the PheWAS, as described above. The methodology used here was identical to our previously published PheWAS on 281,104 UKB participants^10^.

#### P-value threshold

We defined the study-wide significance threshold as p<1×10^-8^. We have previously shown using an n-of-1 permutation approach and the empirical null synonymous model that this threshold corresponds to a false positive rate of 9 and 2, respectively, out of ∼346.5 million tests for binary traits in the setting of collapsing analysis PheWAS^10^.

### Replication analyses

We performed replication analysis of the association between *MAP3K15* and diabetes using the publicly available results from the GWAS in the FinnGen cohort. We accessed the association statistics for the phenome-wide analysis of non-synonymous variants within *MAP3K15* through the FinnGen portal (release 5).

### Secondary association analyses

A total of 40 unique PTVs in *MAP3K15* were observed among the hemizygous male carriers. Two of these PTVs (Arg1122* and Arg1136*) were relatively more frequent. We excluded carriers of these two alleles and re-performed the collapsing analyses for the remaining *MAP3K15* PTVs: Fisher’s exact test for diabetes (‘*20002#1220#diabetes*’) and linear regression for HbA1c.

To determine whether the effect of complete loss of *MAP3K15* on diabetes is mediated via adiposity or the insulin resistance pathway, we performed additional analyses in which we regressed HbA1c and the diabetes phenotype (‘*20002#1220#diabetes*’) on *MAP3K15* PTV carrier status in males, with BMI as the covariate.

To investigate the joint effects of complete loss of *MAP3K15* and a nearby significantly associated indel in *PDHA1* (X-19360844-AAC-A), a gene that overlaps the 3’-UTR of *MAP3K15*, we regressed HbA1c and the diabetes phenotype (‘*20002#1220#diabetes*’) on the carrier status for the two frequent *MAP3K15* PTVs (Arg1122* and Arg1136*) and the *PDHA1* indel in males.

### Expression analyses

We studied previously published bulk RNA-sequencing data available from a mouse insulinoma cell line (β-TC-6) transfected with three different clones carrying MODY-associated variants in NKX6-125. We extracted the DESeq2-derived log fold changes, p-values, and FDR values from the supplementary data. We determined tissue expression using the GTEx portal (http://gtexportal.org/home/). For single-cell RNA-sequencing analysis, we examined eight previously published datasets using tissue from human pancreatic islets spanning 27 healthy donors, five technologies, and four laboratories^17–21^. Data was integrated using Seurat, as previously described^38^.

### Gene-SCOUT

The tool Gene-SCOUT^24^ estimates similarity between genes by leveraging association statistics from the collapsing analysis across 1,419 quantitative traits available in the UKB. We utilised this tool to identify genes that were most similar to the ‘seed gene’ *MAP3K15*.

### Mantis-ML

Mantis-ML^25^ is a gene prioritisation machine learning framework, integrating a diverse set of annotations, including intolerance to variation, tissue expression and animal models. We used this tool to obtain the top disease predictions for *MAP3K15* across 2,536 diseases parsed from Open Targets.

## Data Availability

All data produced in the present work are contained in the manuscript

## Competing Interests

A.N., R.S.D., A.R.H, D.V., A.A., B.B., K.M., B.Z., Q.W., K.S., D.S., B.C., D.S.P., MB., M.S.,

D.B., R.F., M.N.P., and S.P are current employees and/or stockholders of AstraZeneca.

