## Supplementary Information for "Human genetic evidence supports MAP3K15 inhibition as a therapeutic strategy for diabetes"

***Supplemental Information***

Abhishek Nag^1^*, Ryan S. Dhindsa^2^*, Andrew R. Harper^1^, Dimitrios Vitsios^1^, Andrea Ahnmark^3^, Bilada Bilican^4^, Katja Madeyski-Bengtson^4^, Bader Zarrouki^3^, Quanli Wang^2^, Katherine Smith^1^, Dave Smith^5^, Benjamin Challis^6^, Dirk S. Paul^1^, Mohammad Bohlooly-Y^4^, Mike Snowden^7^, David Baker^8^, Regina Fritsche-Danielson^9^, Menelas N. Pangalos^10^, Slavé Petrovski^1,11^

^1^Centre for Genomics Research, Discovery Sciences, BioPharmaceuticals R&D, AstraZeneca, Cambridge, UK

^2^Centre for Genomics Research, Discovery Sciences, BioPharmaceuticals R&D, AstraZeneca, Waltham, USA

^3^Bioscience Metabolism, Early CVRM, BioPharmaceuticals R&D, AstraZeneca, Gothenburg, Sweden

^4^Discovery Biology, Discovery Sciences, BioPharmaceuticals R&D, AstraZeneca, Gothenburg, Sweden

^5^Emerging Innovations, Discovery Sciences, BioPharmaceuticals R&D, AstraZeneca, Cambridge, UK

^6^Translational Science and Experimental Medicine, Early CVRM, BioPharmaceuticals R&D, AstraZeneca, Cambridge, UK

^7^Discovery Sciences, BioPharmaceuticals R&D, AstraZeneca, Cambridge, UK

^8^Bioscience Metabolism, Early CVRM, BioPharmaceuticals R&D, AstraZeneca, Cambridge, UK

^9^Early CVRM, BioPharmaceuticals R&D, AstraZeneca, Gothenburg, Sweden

^10^BioPharmaceuticals R&D, AstraZeneca, Cambridge, UK

^11^Departments of Medicine and Neurology, University of Melbourne, Royal Melbourne Hospital, Melbourne, Victoria, Australia

*These authors contributed equally

**Corresponding author:**
Slavé Petrovski

Vice-President, Centre for Genomics Research,

Discovery Sciences, BioPharmaceuticals R&D
AstraZeneca
Cambridge

United Kingdom


**Supplementary Tables**

**Supplementary Table 1: The diabetes-related clinical phenotypes and quantitative traits from the UK Biobank analysed in this study**

Provided as separate additional file

Supplementary Table 1A: The diabetes-related clinical phenotypes from the UK Biobank were captured using the ICD-10 codes E10-E14 and any additional phenotypic terms containing the strings ‘diabetes’ or ‘Diabetes’. Phenotypic terms related to diabetes insipidus, diabetes medications, or other endocrine conditions were excluded. We additionally performed manual inspection of the 90 diabetes-related clinical phenotypes that were selected for this study.

Phenotypes with less than 5 cases in a particular ancestry were not analysed and the corresponding case/control numbers have been labelled as “NA”.

Supplementary Table 1B*:* Five quantitative traits related to diabetes were analysed for genes that were significantly associated with one of the diabetes-related clinical phenotypes.

**Supplementary Table 2: Summary of the different models implemented in the gene-level collapsing analysis.**

On next page

Criteria used for determining qualifying variants (QVs) for the 10 different nonsynonymous models used in the gene-level collapsing analysis. In addition, a synonymous collapsing model was used for the purpose of establishing an empirical negative control.

| **Collapsing model** | **gnomAD  MAF*** | **UKB  MAF** | **UKB cohort  no call or QC fail^** | **Variant  type** | **REVEL**  **cut-off** | **Missense tolerance ratio (MTR) cut-offs** |
| --- | --- | --- | --- | --- | --- | --- |
| **syn (synonymous negative control)** | ≤0.005% | ≤0.05% | ≤0.005% | Synonymous | - | - |
| **ptv      (Protein Truncating)** | ≤0.1% (popmax) | ≤0.1% | ≤0.01% | PTV | - | - |
| **ptv5pcnt (Protein Truncating, ≤5% MAF)** | ≤5% (popmax) | ≤5% | ≤0.5% | PTV | - | - |
| **UR      (Ultra-rare damaging)** | 0% | ≤0.005% | ≤0.001% | Non-synonymous | ≥0.25 | - |
| **URmtr (Ultra-rare damaging, MTR informed)** | 0% | ≤0.005% | ≤0.001% | Non-synonymous | ≥0.25 | MTR≤25^th^ %ile or intragenic MTR≤50^th^ %ile |
| **raredmg (Rare damaging)** | ≤0.005% | ≤0.025% | ≤0.005% | Missense | ≥0.25 | - |
| **raredmgmtr (Rare damaging, MTR informed)** | ≤0.005% | ≤0.025% | ≤0.005% | Missense | ≥0.25 | MTR≤25^th^ %ile or  intragenic MTR≤50^th^ %ile |
| **flexdmg (Flexible MAF, damaging non-synonymous)** | ≤0.1% (popmax) | ≤0.1% | ≤0.01% | Non-synonymous | ≥0.25 | - |
| **flexnonsynmtr (Flexible MAF, non-synonymous, MTR informed)** | ≤0.1% (popmax) | ≤0.1% | ≤0.01% | Non-synonymous | - | MTR≤25^th^ %ile or  intragenic MTR≤50^th^ %ile |
| **ptvraredmg (PTV or rare damaging models combined)** | PTV≤0.1% (popmax)  missense≤0.005%  and ≤0.05% (popmax) | PTV≤0.1%, missense≤ 0.025% | ≤0.01% | Non-synonymous | ≥0.25 | - |
| **rec (Non-synonymous recessive)** | ≤1% (popmax)  ≤10 homozygous calls | ≤1% | ≤0.1% | Non-synonymous | - | - |

(MAF = Minor Allele Frequency; QC = Quality Control; MTR = Missense Tolerance Ratio)
*reflects the gnomAD global_raw MAF unless otherwise specified. 
^reflects the maximum proportion of UKB exome sequences permitted to either have ≤ 10-fold coverage at variant site or carry a low-confidence variant that did not meet one of the quality-control thresholds applied to collapsing analyses (see methods). 
**Synonymous**: synonymous_variant 
**PTV**: exon_loss_variant, frameshift_variant, start_lost, stop_gained, stop_lost, splice_acceptor_variant, 
splice_donor_variant, gene_fusion, bidirectional_gene_fusion, rare_amino_acid_variant, transcript_ablation 
**Missense**: missense_variant_splice_region_variant, missense_variant 
**Nonsynonymous**:exon_loss_variant, frameshift_variant, start_lost, stop_gained, stop_lost, splice_acceptor_variant,splice_donor_variant, gene_fusion, bidirectional_gene_fusion, rare_amino_acid_variant, transcript_ablation, 
conservative_inframe_deletion, conservative_inframe_insertion, disruptive_inframe_insertion, 
disruptive_inframe_deletion, missense_variant_splice_region_variant, missense_variant, protein_altering_variant

**Supplementary Table 3: Distinct gene-phenotype relationships identified for the four genes that were significantly associated with at least one diabetes-related clinical phenotypes in the gene-level collapsing analysis among European ancestry participants in the UK Biobank**

Provided as separate additional file

The diabetes-related clinical phenotypes that were associated (p<1x10^-7^) with each of the four genes significantly associated (p<1x10^-8^) with at least one diabetes-related clinical phenotype in the gene-level collapsing analysis among European ancestry participants in the UK Biobank. The most significant collapsing model and the corresponding association statistics have been provided for each gene-phenotype relationship.

(Chr=Chromosome, QV=Qualifying Variant, OR=Odds Ratio, CI=Confidence Intervals)

**Supplementary Table 4: Genes significantly associated with diabetes-related clinical phenotypes in the pan-ancestry gene-level collapsing analysis in the UK Biobank**

Provided as separate additional file

A pan-ancestry was performed by combining results from the gene-level collapsing analyses for the four major ancestral groups in the UKB (Europeans, Africans, South Asians and East Asians) via a Cochran-Mantel-Haenszel test. The most significant diabetes-related clinical phenotype and the corresponding association statistics have been provided for the seven genes that were significantly associated (p<1x10^-8^) with at least one diabetes-related clinical phenotype.

(Chr=Chromosome, QV=Qualifying Variant, OR=Odds Ratio, CI=Confidence Intervals, CMH= Cochran-Mantel-Haenszel test)

**Supplementary Table 5: Effect of complete loss of *MAP3K15* on diabetes-related traits**

Provided as separate additional file

The effect of complete loss of *MAP3K15* on HbA1c and diabetes-related clinical phenotypes was evaluated by comparing hemizygous male PTV carriers to male non-carriers of European ancestry. The diabetes-related clinical phenotypes that were associated with *MAP3K15* in the initial collapsing analysis were selected.

(OR=Odds Ratio, CI=	Confidence Intervals)

**Supplementary Table 6: Protein-truncating variants in *MAP3K15* observed among the European ancestry males in the UK Biobank**

Provided as separate additional file

Hemizygous carriers for 40 unique protein-truncating variants in *MAP3K15* were observed among the European ancestry males in the UK Biobank. Two particular protein-truncating variants (Arg1122* and Arg1136*), that were relatively more frequent among the carriers, have been highlighted.

(MAF=Minor Allele Frequency)

**Supplementary Table 7: Effects of the *MAP3K15* PTVs and the *PDHA1* indel on diabetes-related traits**

Provided as separate additional file

The effects of the two relatively more frequent *MAP3K15* PTVs [Arg1122* and Arg1136*] and the collection of remaining PTVs on HbA1c and diabetes were each tested separately.

An indel (X-19360844-AAC-A) in the 3’-UTR of *PDHA1*, which overlaps with *MAP3K15*, is also significantly associated with HbA1c. The joint effects of the two *MAP3K15* PTVs and the *PDHA1* indel were tested.

(OR=Odds Ratio, CI=Confidence Intervals)

**Supplementary Table 8: *MAP3K15* associations with diabetes-related clinical phenotypes in the FinnGen cohort**

Provided as separate additional file

Association statistics for variants in *MAP3K15* that were associated with type 1 or type 2 diabetes in the FinnGen cohort have been provided.

(OR=Odds Ratio)

**Supplementary Table 9: Non-diabetes-related clinical phenotypes associated with *MAP3K15* in the collapsing analysis across all ancestries in the UK Biobank**

Provided as separate additional file

Supplementary Table 9A: A total of 10 non-diabetes-related traits were suggestively associated (p<1x10^-4^) with *MAP3K15* in the collapsing analysis across all non-synonymous models and ancestries in the UK Biobank.

Supplementary Table 9B: Association of hypertension-related traits (having at least one case QV carrier) with recessive form of *MAP3K15* missense variants (”recmissense” model) and PTVs (“recptv” model) in the UK Biobank.

(QV=Qualifying Variants, OR=Odds Ratio, CI=Confidence Intervals)

**Supplementary Table 10: Association of complete loss of *MAP3K15* with blood pressure-related traits among European ancestry participants in the UK Biobank**

Provided as separate additional file

The effect of complete loss of *MAP3K15* in hemizygous male PTV carriers on blood pressure-related traits was tested. Quantitative traits that were analysed included automated measurements of systolic and diastolic blood pressure (UK Field IDs: 4080 and 4079, respectively). Binary traits that were analysed included a diagnosis of hypertension (*Union#I10#I10 Essential (primary) hypertension*) and a specific hypertensive phenotype that showed suggestive association in the South Asian ancestry participants (*41202#BlockI10-I15#I10-I15 Hypertensive diseases*).

(OR=Odds Ratio, CI=Confidence Intervals)

**Supplementary table 11: Mantis-ML predictions of *MAP3K15* disease associations**

Provided as separate additional file

The list provides the Mantis-ML predictions (‘Disease rank’ and ‘Gene rank percentile’) for *MAP3K15* for the entire set of 2,536 diseases that were parsed from Open Targets.

**Supplementary Table 12: Commonly prescribed blood pressure-lowering medications in the UK Biobank**

Provided as separate additional file

The list provides the number of participants taking each of the commonly prescribed blood pressure-lowering medications in the UK Biobank. Analyses involving blood pressure measurements (systolic and diastolic blood pressure) were adjusted for the intake of one of these blood pressure-lowering medications.

**Supplementary Figures**

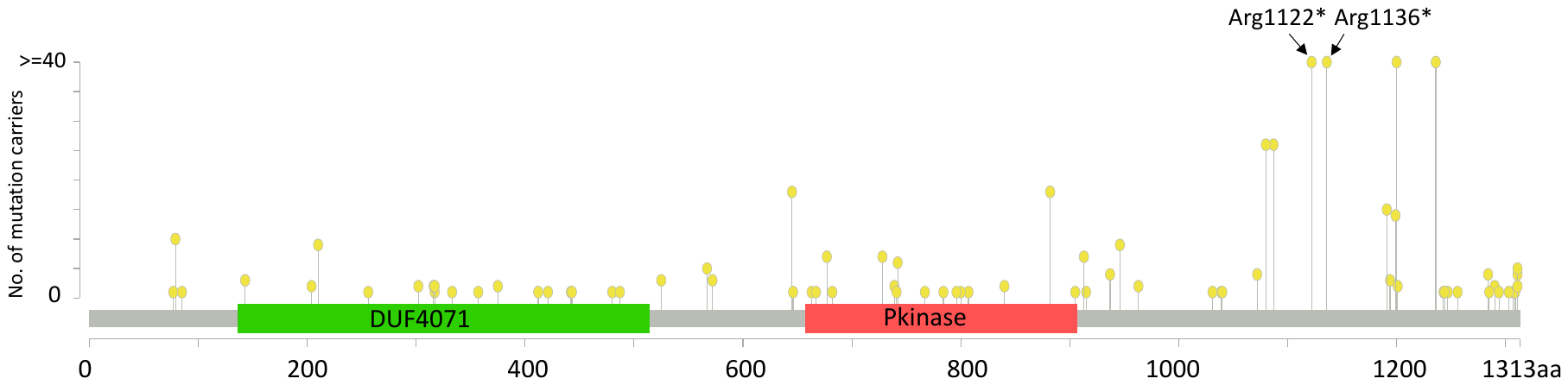

**Supplementary Figure 1: Distribution of PTVs in the *MAP3K15* gene sequence across all ancestries in the UK Biobank**

Lollipop plot depicting *MAP3K15* PTVs (stop gain and frameshift variants) observed among participants across all ancestries in the UK Biobank. Essential splice variants were not included in the lollipop plot. The two most frequent PTVs have been annotated. Just one individual in the entire cohort carried both PTVs. The y-axis was capped at 40.

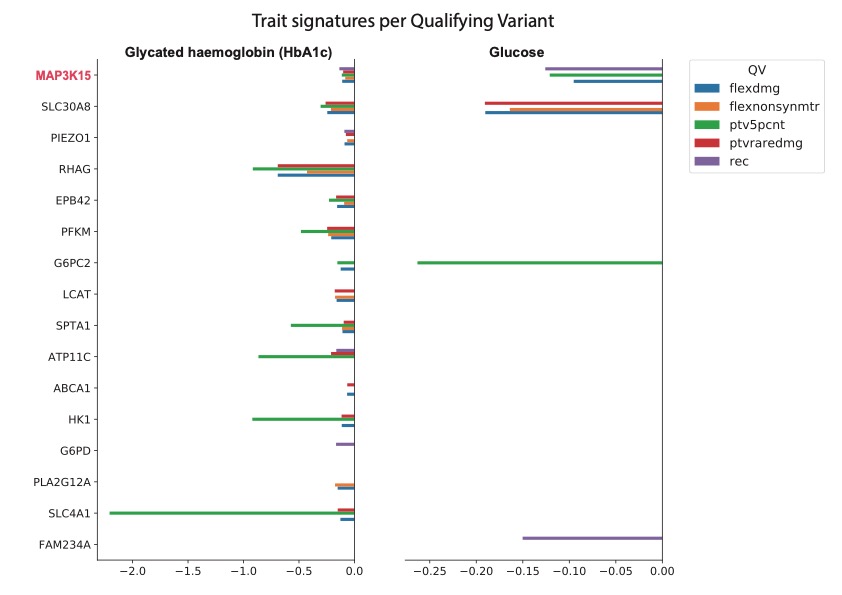

**Supplementary Figure 2: Association signatures with HbA1c and glucose for genes most similar to *MAP3K15***.

Comparison of linear regression coefficients for HbA1c and glucose between *MAP3K15* and the genes most “similar” to it, derived from Gene-SCOUT. All collapsing models for which *MAP3K15* showed significant associations with HbA1c and glucose have been provided.

**Supplemental Methods**

**Joint analysis of the *MAP3K15* and *PDHA1* loci**

*MAP3K15* overlaps with the 3’-UTR of *PDHA1,* a gene that encodes a subunit of the enzyme pyruvate dehydrogenase and catalyzes a step in the glycolysis pathway. Moreover, an indel (X-19360844-AAC-A) in the 3’-UTR of *PDHA1* is significantly associated with HbA1c levels in the UKB (beta = -0.13, 95% CI: [-0.17, -0.09], *P* = 2.1x10^-11^). We performed a conditional analysis to ensure that the observed effect of the two more common *MAP3K15* PTVs on HbA1C levels was independent of the *PDHA1* indel. Indeed, the associations for all three variants with HbA1c remained significant in the joint analysis (**Supplementary Table 7**).
